# Microtesla Magnetic Therapy for cognitive impairment in post-acute sequelae of SARS CoV-2: A randomized controlled feasibility study

**DOI:** 10.64898/2026.03.18.26348530

**Authors:** Alexandra Canori, Eric Watson, Devanshi Patel, Arianna Fiorentino, Christopher Santiago, David Maltz, Blake Gurfein, David Putrino, Jacqueline Becker

**Author notes:** Corresponding author: David Putrino, PhD, 5 E 98^th^ St SB-18, New York, NY 10029. Authors contributed equally to this work (Alexandra Canori, Eric Watson, David Putrino, Jacqueline Becker).

## Abstract

**Background:** Cognitive impairment has significant implications for function and quality of life and is common in individuals with post-acute sequelae of SARS CoV-2, also known as long COVID (LC). Emerging evidence suggests that sustained neuroinflammation, cerebrovascular dysfunction, and mitochondrial impairment are contributors to cognitive symptoms. Microtesla Magnetic Therapy (MMT) is a low-amplitude radiofrequency magnetic field intervention that has demonstrated anti-inflammatory and neuroprotective effects in preclinical models, suggesting it may be valuable in the management of cognitive impairment from LC and other neurological disorders. This study is the first randomized controlled trial to evaluate MMT for LC-related cognitive impairment.

**Objective:** To evaluate the feasibility, safety, and preliminary efficacy of an at-home MMT intervention in individuals with moderate-to-severe cognitive impairment from LC.

**Methods:** In this prospective feasibility study, 30 participants with LC-related cognitive impairment were randomized (2:1) to receive active or sham MMT. Participants self-administered 15-minute treatments at home with remote monitoring twice weekly for 4 weeks using a head-worn device that delivered a nonthermal radiofrequency magnetic field to the whole brain. Feasibility was defined as completion of at least 80% of prescribed treatments and all study visits. Secondary outcomes included safety, cognitive function, and self-reported mood and quality of life assessed at baseline, post-treatment (Week 4), and follow-up (Week 8).

**Results:** Feasibility was high, with 100% treatment adherence among participants who completed the study and strong usability ratings for at-home administration. There were no device-related adverse events. Compared with sham, participants receiving active MMT showed significantly greater improvements from baseline to Week 8 in WAIS-IV Digit Span Sequencing (p= 0.026), HVLT-R Recall (p= 0.044), and D-KEFS Color Naming (p= 0.049). Additional measures of attention, processing speed, and executive function demonstrated favorable trends in the active group. Emotional well-being, assessed by the SF-36, improved significantly in the active group at Week 8 compared with sham (p= 0.017), and mood symptoms showed clinically meaningful improvement.

**Conclusions:** Administration of the MMT intervention at home was feasible, safe, and well tolerated in individuals with cognitive impairment from LC. Preliminary findings suggest sustained clinically meaningful improvements in multiple cognitive domains and mood following treatment.

**Trial Registration:** ClinicalTrials.gov NCT06739668, https://clinicaltrials.gov/study/NCT06739668, 2024-12-17

## Introduction

Approximately 10-26% of patients infected with SARS-CoV-2 develop post-acute sequelae of COVID-19, or long COVID (LC),^1^ a condition characterized by symptoms including fatigue, mood changes, sleep disruption, and cognitive impairment that persist at least 3 months following acute infection.^2^ CDC estimates LC affects 6.4% of adults in the United States, which is approximately 17 million individuals.^3^ Multiple pathophysiologic mechanisms have been implicated in the development of LC, such as persistent peripheral and central nervous system immune activation, incomplete viral clearance or latent viral reservoirs, immune cell exhaustion, vascular and endothelial injury, gut dysbiosis, autoantibody production, and dysregulated autonomic signaling.^4–6^ The societal burden of LC is substantial, with estimated annual costs of $2.6 trillion in the United States alone due to lost productivity, increased healthcare utilization, and reduced quality of life.^7^

Cognitive impairment is among the most prevalent and disabling features of LC, with estimates suggesting that 20-30%, or approximately 3-5 million, individuals with LC experience persistent cognitive difficulties.^8,9^ Emerging evidence implicates neuroinflammation as a key underlying driver of cognitive impairment.^10,11^ Specifically, neuroimmune activation and subsequent oxidative stress may perturb mitochondrial bioenergetics and promote a self-perpetuating cycle of energy failure and chronic neuroinflammation, a plausible substrate for the persistent “brain fog” reported by patients.^12^ Supporting this framework, several neuroimaging studies have demonstrated microglial activation and elevated inflammatory markers in individuals with LC-related cognitive symptoms.^13,14^ The neurocognitive profile in LC is particularly notable for deficits in executive functioning, attention, and processing speed,^15^ which appear to persist over time.^16^ Despite growing mechanistic insight, no therapeutic interventions have demonstrated efficacy in targeting both neuroinflammation and mitochondrial dysfunction.

Low-amplitude magnetic field interventions have demonstrated safety and tolerability, with evidence of rapid anti-inflammatory effects in post-surgical clinical studies^17^ and animal models of neuroinflammation.^18^ Building on this foundation, Microtesla Magnetic Therapy (MMT) was developed as a non-invasive approach to deliver carefully tuned radiofrequency magnetic fields to the brain. Recent preclinical studies showed that MMT suppresses NF-κB signaling, reduces neuronal reactive oxygen species, and decreases glial activation and lesion burden *in vivo*, producing sustained anti-inflammatory and neuroprotective effects.^19^ These findings suggest MMT engages redox-sensitive inflammatory networks and modulates neuroimmune activation, pathways increasingly recognized as central drivers of cognitive dysfunction. As a result, MMT represents a promising nonpharmacologic strategy to address a significant unmet clinical need by directly targeting neuroinflammatory biology.

Based upon the noninvasive, low-amplitude nature of MMT and compelling mechanistic evidence, the present study evaluated the feasibility of at-home MMT as a therapeutic intervention for individuals with moderate to severe cognitive impairment from LC. Our objectives were to (1) assess treatment feasibility and acceptability in this population, (2) evaluate the safety and tolerability, and (3) collect preliminary efficacy data to inform the design of larger clinical trials. We hypothesized that MMT would be feasible and safe in individuals with moderate-to-severe cognitive impairment from LC, and that active treatment would demonstrate greater improvements in cognitive function and mood symptoms compared to sham treatment.

## Methods

### Study Design

A prospective, triple-blind, randomized controlled trial of 30 participants with LC and moderate-to-severe cognitive impairment was conducted to evaluate the feasibility, safety, and preliminary efficacy of MMT. Participants were randomized in a 2:1 ratio to receive either an active or sham device. Participants, investigators, and outcome assessors were blinded to group allocation.

Device assignment was performed by an independent study engineer who pre-programmed active and sham devices prior to delivery to the study site, ensuring allocation concealment. The study protocol was approved by the Program for the Protection of Human Subjects at the Icahn School of Medicine at Mount Sinai (STUDY-24-01276) and registered on ClinicalTrials.gov (NCT06739668). All participants provided written informed consent prior to participation.

### Study Participants

The following inclusion criteria were used to determine subject eligibility: (1) Age 18 years or older, (2) English speaking, (3) SARS CoV-2 infection documented by laboratory nucleic acid amplification test or antibody test ≥6 months from screening, (4) Experiencing symptoms ≥6 months that meet LC diagnostic criteria,^2^ (5) Objective cognitive impairment (defined by ≤1 standard deviations below the normative mean) on at least one measure of executive functioning, (6) Individuals of childbearing age agreeing to use a highly effective form of birth control, and (7) Willing and able to provide informed consent and attend all study visits.

Exclusion criteria were determined by self-report or review of electronic health record, and included: pre-existing cognitive impairment, current use of immunomodulatory medications, active psychiatric or neurological disorders, cranially implanted devices or pacemakers, severe head injury within 12 months, SARS-CoV-2 reinfection or vaccination within 30 days, diagnosis of immune/autoimmune condition prior to SARS-CoV-2 infection, change in anti-depressant or other psychoactive medication within 90 days, BrainCheck assessment within the last 6 months, febrile at the time of enrollment visit, enrollment in another clinical trial within 90 days of study period, inability to achieve appropriate positioning of the study device, and pregnancy.

### Intervention

Participants randomized to the active group received treatment with the MMT device, which delivers a low-amplitude nonthermal radiofrequency magnetic field at 27.12 MHz via a head-worn applicator (Figure 1). Sham devices were visually identical and had the same indicator lights and sounds, but emitted no magnetic field. Participants were trained to self-administer treatments at home twice weekly for 15-minute sessions, separated by at least 72 hours, a time interval informed by previous *in vivo* studies.

**Figure 1.**
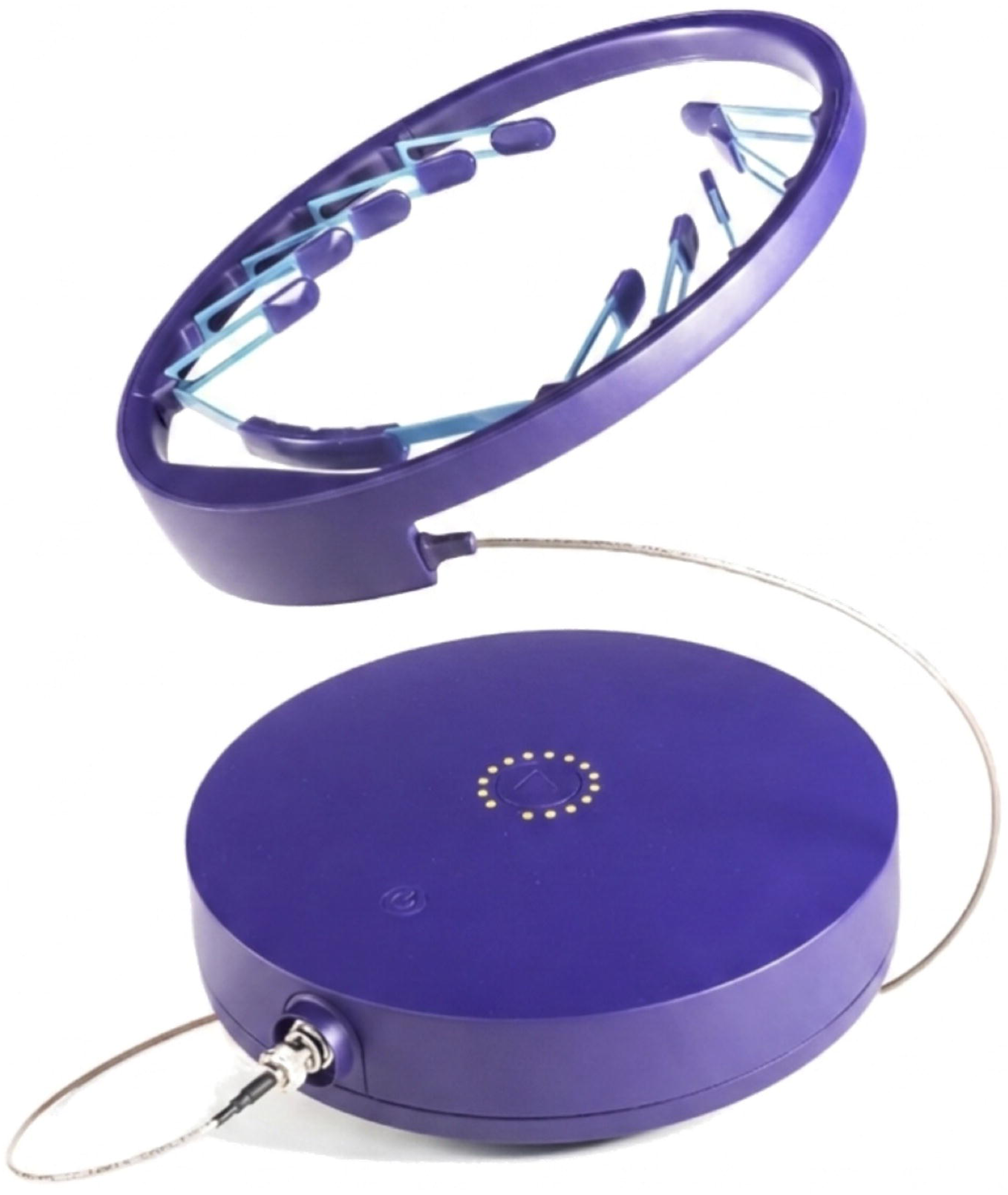
Mity Biophysics Delivery System

### Study Outcomes

The primary outcome was feasibility, defined as the proportion of participants completing at least 80% of scheduled treatments and all three study visits. Additional feasibility measures included Device Usability Ratings (Supplementary Table 1) and completeness of data collection.

Secondary outcomes included safety, cognitive performance, and self-reported symptom changes. Safety outcomes were assessed weekly via REDCap questionnaires and through in-person evaluations at study visits. Adverse events were recorded and graded according to ISO 14155 criteria.

Objective cognitive performance was assessed with a standardized neurocognitive battery across domains, including attention, processing speed, language, memory, and executive functioning.

Measures included the Wechsler Adult Intelligence Scale Fourth Edition (WAIS-IV) Digit Span, Hopkins Verbal Learning Test–Revised (HVLT-R), Delis-Kaplan Executive Function System (D-KEFS) Color-Word Interference Test, DKEFS Verbal Fluency Test, Trail Making Test (TMT) Parts A & B, Ruff 2 & 7, Brief Visuospatial Memory Test (BVMT), Rey Complex Figure Test (RCFT), Symbol Digit Modalities Test (SDMT), and Multilingual Naming Test (MINT).

All assessments were administered in a fixed order by trained staff to minimize order effects and provide consistency across participants.

Mood and quality of life outcomes were assessed using standardized instruments including the Patient Health Questionnaire 9 (PHQ-9), Generalized Anxiety Disorder 7 (GAD-7), DePaul Symptom Questionnaire Post-Exertional Malaise (DSQ-PEM), PROMIS Fatigue Short Form-7a, and 36-Item Short Form Health Survey (SF-36). A description of each of these measures is provided in Table 1.

**Table 1.**
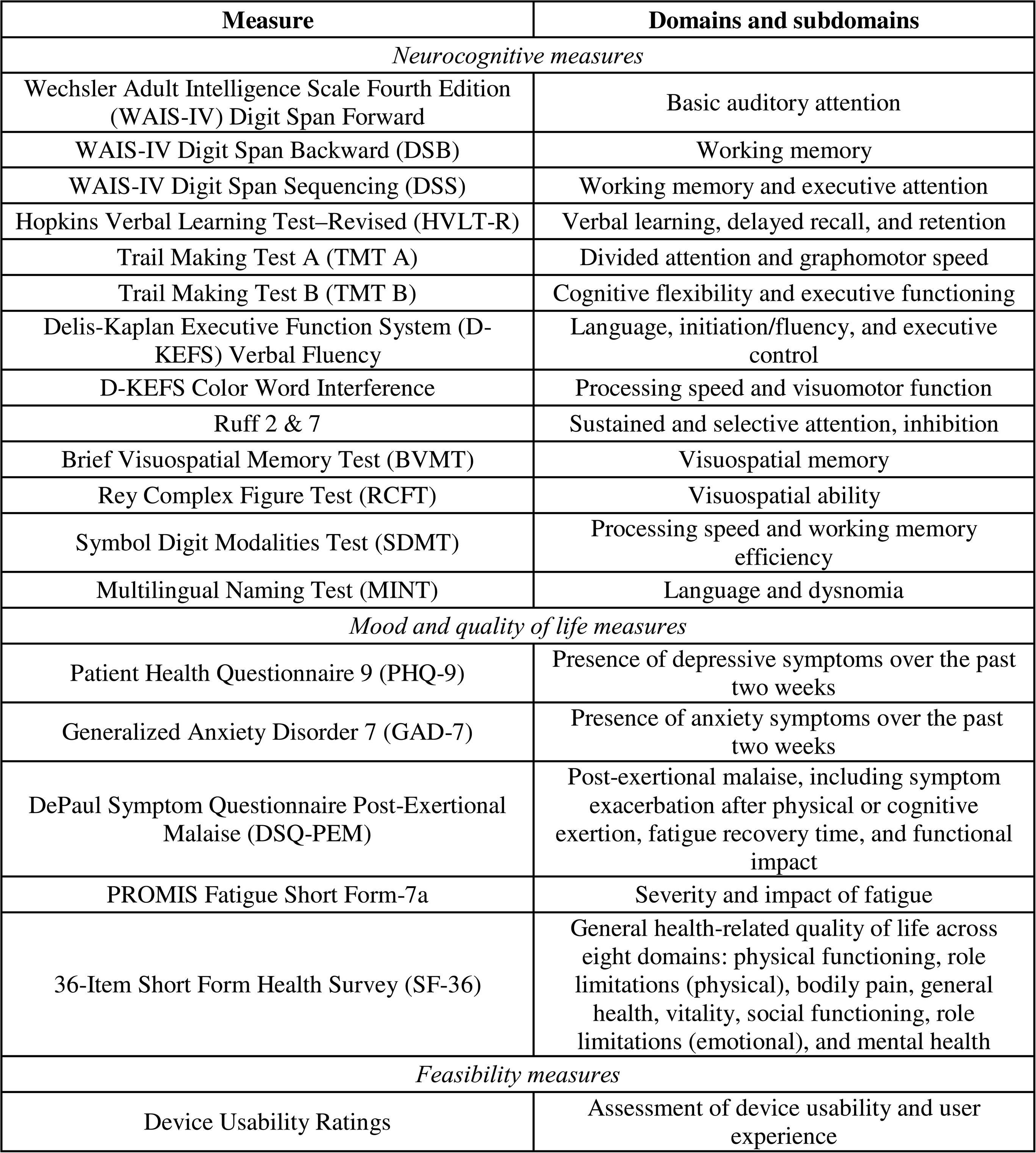
Outcome measures and description of domains assessed.

| Measure | Domains and subdomains |
| --- | --- |
| *Neurocognitive measures* |  |
| Wechsler Adult Intelligence Scale Fourth Edition (WAIS-IV) Digit Span Forward | Basic auditory attention |
| WAIS-IV Digit Span Backward (DSB) | Working memory |
| WAIS-IV Digit Span Sequencing (DSS) | Working memory and executive attention |
| Hopkins Verbal Learning Test–Revised (HVLTR) | Verbal learning, delayed recall, and retention |
| Trail Making Test A (TMT A) | Divided attention and graphomotor speed |
| Trail Making Test B (TMT B) | Cognitive flexibility and executive functioning |
| Delis-Kaplan Executive Function System (D-KEFS) Verbal Fluency | Language, initiation/fluency, and executive control |
| D-KEFS Color Word Interference | Processing speed and visuomotor function |
| Ruff 2 & 7 | Sustained and selective attention, inhibition |
| Brief Visuospatial Memory Test (BVMT) | Visuospatial memory |
| Rey Complex Figure Test (RCFT) | Visuospatial ability |
| Symbol Digit Modalities Test (SDMT) | Processing speed and working memory efficiency |
| Multilingual Naming Test (MINT) | Language and dysnomia |
| *Mood and quality of life measures* |  |
| Patient Health Questionnaire 9 (PHQ-9) | Presence of depressive symptoms over the past two weeks |
| Generalized Anxiety Disorder 7 (GAD-7) | Presence of anxiety symptoms over the past two weeks |
| DePaul Symptom Questionnaire Post-Exertional Malaise (DSQ-PEM) | Post-exertional malaise, including symptom exacerbation after physical or cognitive exertion, fatigue recovery time, and functional impact |
| PROMIS Fatigue Short Form-7a | Severity and impact of fatigue |
| 36-Item Short Form Health Survey (SF-36) | General health-related quality of life across eight domains: physical functioning, role limitations (physical), bodily pain, general health, vitality, social functioning, role limitations (emotional), and mental health |
| *Feasibility measures* |  |
| Device Usability Ratings | Assessment of device usability and user experience |

### Study Procedures

Participants attended three in-person visits (Week 0, Week 4, Week 8). At Visit 1, participants completed informed consent, medical history review, and baseline cognitive and self-report assessments. Participants were trained to use the study device and self-administered the first 15-minute treatment under supervision of clinical staff. They were instructed to continue treatments twice per week at home, separated by at least 72 hours for the 4-week duration of the treatment period. Each treatment was scheduled as a video visit with the study team, and a team member visually confirmed the accurate positioning of the device and administration of the treatment.

The device automatically entered a 72-hour lockout period after each treatment to prevent deviations from the treatment protocol. Participants returned to the study site for Visits 2 and 3 and repeated all assessments conducted at baseline to evaluate treatment effects and post-treatment persistence. All visits were scheduled at the same time of day to reduce diurnal variability in performance measures.

### Data Management

The study was monitored by the principal investigator and study staff. All data were entered into a 21 CFR 11-compliant electronic data capture system. Treatment adherence, device logs, and questionnaire completion were monitored weekly, and self-report safety questionnaires were administered weekly to participants during the treatment period and reviewed by the study team. Data quality checks were performed regularly to ensure accuracy and completeness.

### Data Sharing Statement

De-identified data and supporting documents including study protocol and statistical analysis plan will be available upon reasonable request. Requests must be approved by the corresponding author.

### Statistical Analysis

#### Feasibility and Safety Analyses

Feasibility was assessed by the proportion of participants completing at least 80% of prescribed treatments and all scheduled visits. Device usability ratings and responses were summarized. All adverse events and complications occurring during the treatment and follow-up periods were documented and summarized by treatment group.

#### Neurocognitive and Self-Report Measures

Neurocognitive instruments and self-report questionnaires were used to evaluate within-subject and between-group changes from baseline (week 0) to post-treatment (week 4) and from baseline to follow-up (week 8). The study was exploratory and not statistically powered to detect prespecified effect sizes for each endpoint. Clinically meaningful change for neurocognitive assessments of cognitive function was set to 0.5 standard deviations.^20^ Clinically meaningful change for PHQ and GAD-7 was interpreted as 2 points, aligning with previous studies.^21^ Descriptive statistics are reported as means and standard deviations (SD) for continuous variables, and as counts and percentages for categorical variables. All statistical tests are two-sided, with an alpha level of 0.05 used to determine significance. No p-value adjustments were made for multiple comparisons; however, confidence intervals are provided for further assessment of the observed differences. Cohen’s d was calculated to measure effect size in select measures and are reported in Supplementary Table 2. Analyses were conducted using complete-case data, imputation was not used for missing values. Statistical analysis was performed using JMP Pro software (version 19.0).

## Results

### Participant demographics

A total of 65 participants were screened; 32 participants failed screening, and 33 subjects were enrolled (Figure 2). To preserve the randomization allocation and ensure that 30 participants completed all study procedures, screening exceeded the target sample size to account for anticipated attrition. Three study participants were withdrawn from the study after enrollment for the following reasons: (1) COVID reinfection during the study period, which per protocol required withdrawal; (2) Two consecutive video visits missed, which per protocol required withdrawal; (3) Participant injured their ankle and withdrew to pursue surgical intervention. In total, 30 participants with LC-related cognitive impairment completed all trial activities (active= 20; sham= 10). Groups were comparable at baseline with respect to age, education, sex, ethnicity, and time since SARS-CoV-2 infection (Table 2). Differences in race were observed between groups; however, subgroup sizes were small, limiting interpretability.

**Figure 2.**
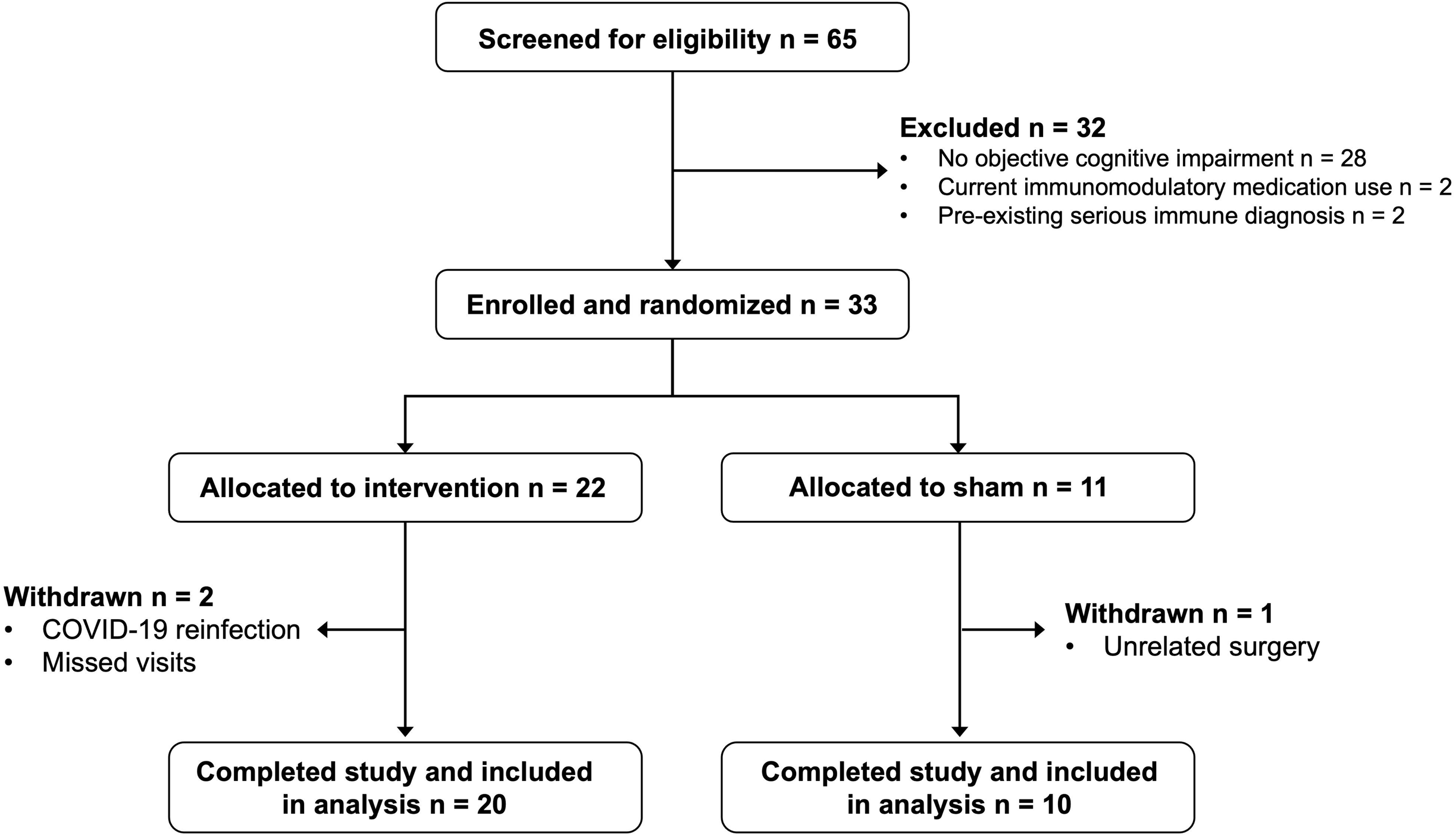
CONSORT Flow Diagram

**Table 2.** Baseline Demographic and Clinical Characteristics of Study Participants.

| **Variable** |  | **Treatment** |  |  | **P-value** |
| --- | --- | --- | --- | --- | --- |
| **Variable** |  | **Sham (N = 10)** | **Active (N = 20)** | **All (N = 30)** |  |
| Age |  | 48.5 (11.7) | 47.7 (13.9) | 47.9 (13.0) | 0.870 t-test |
| Sex | Female | 8 (80.0%) | 13 (65.0%) | 21 (70.0%) | 0.675 Fisher's exact |
|  | Male | 2 (20.0%) | 7 (35.0%) | 9 (30.0%) |  |
| Education (in years) |  | 17.9 (2.7) | 17.5 (2.4) | 17.6 (2.5) | 0.648 t-test |
| Race | Asian | 1 (10.0%) | 2 (10.0%) | 3 (10.0%) | 0.011 Fisher's exact |
|  | Black | 2 (20.0%) | 1 (5.0%) | 3 (10.0%) |  |
|  | White | 3 (30.0%) | 16 (80.0%) | 19 (63.3%) |  |
|  | Other | 3 (30.0%) | 0 (00.0%) | 3 (10.0%) |  |
| Ethnicity | Hispanic or Latino | 2 (20.0%) | 3 (15.0%) | 5 (16.7%) | 1.0 Fisher's exact |
|  | Non-Hispanic or Latino | 8 (80.0%) | 17 (85.0%) | 25 (83.3%) |  |
| Months since SARS-CoV-2 infection |  | 38.9 (12.9) | 45.6 (15.7) | 43.4 (15.0) | 0.513 t-test |

### Feasibility and Safety

This study aimed to determine the feasibility of applying the MMT device for treatment at home. All 30 participants who completed the study received all 8 treatments in accordance with the protocol, although one participant who did not complete the study was withdrawn due to missing two consecutive visits. Correct positioning of the device on the head and completion of the full 15-minute treatment duration was confirmed by study staff. Two adverse events occurred in the active treatment group: an ankle injury and SARS-CoV-2 reinfection. Both adverse events were mild and deemed unrelated to treatment. No serious adverse events occurred during this study.

### Neurocognitive Outcomes

Across multiple neurocognitive domains, participants receiving active MMT demonstrated greater improvements than those receiving sham treatment, with effects most evident at the 8-week follow-up (Table 3). Significant improvements in the active treatment with small to medium effect sizes were observed in between-group analysis from baseline to 8 weeks for DSS (d= 0.536, p= 0.026), HVLT-R Recall (d= 0.229, p= 0.044), and D-KEFS Color Naming (d= 0.698), p= 0.049). Additional measures of attention, processing speed, inhibition, and executive function including Ruff 2&7 controlled speed (d= 0.615, p= 0.158), Ruff 2&7 total speed (d= 0.619, p= 0.093), D-KEFS word reading (d= 0.763, p= 0.818), inhibition (p= 0.946), inhibition/switching (p= 0.817), letter fluency (d= 0.327, p= 0.698), category switching (p= 0.624), TMT A (d= 0.664, p= 0.170), TMT B (p= 0.826), Ruff 2&7 automatic speed (d= 0.574, p= 0.076), DSF (d= 0.225, p= 0.780), DSB (d= 0.316, p= 0.247) and BVMT Total Recall (p= 0.666) showed greater improvements in the active group compared with sham, though between-group comparisons did not reach statistical significance in this exploratory sample. The D-KEFS Category Fluency, RCFT, SDMT, and MINT did not show improvements from baseline to 8 weeks (Supplementary Table 3).

**Table 3.** Between-group comparisons of neurocognitive scores and mood and quality of life scores.

| Outcome | Week | Sham Mean (SD) | Active Mean (SD) | Active-Sham Difference (95% CI) | P-value† |
| --- | --- | --- | --- | --- | --- |
| *Neurocognitive Outcomes* |  |  |  |  |  |
| Digit Span Forward | 4 ΔBL | 1.10 (5.63) | 4.30 (7.60) | 3.20 (-1.88, 8.28) | 0.206 |
|  | 8 ΔBL | 3.20 (6.00) | 3.90 (7.11) | 0.70 (-4.45, 5.85) | 0.780 |
| Digit Span Backward | 4 ΔBL | 3.80 (9.67) | 3.00 (7.12) | -0.80 (-8.19, 6.59) | 0.820 |
|  | 8 ΔBL | 2.70 (4.83) | 5.65 (8.83) | 3.00 (-2.17, 8.07) | 0.247 |
| Digit Span Sequencing | 4 ΔBL | -0.50 (4.14) | 1.30 (7.35) | 1.80 (-2.51, 6.11) | 0.399 |
|  | 8 ΔBL | -2.10 (4.12) | 3.30 (8.49) | 5.40 (0.68, 10.12) | **0.026** |
| HVLT-R Total Recall | 4 ΔBL | -4.13 (12.25) | 1.34 (13.95) | 5.47 (-4.89, 15.83) | 0.284 |
|  | 8 ΔBL | -4.38 (9.31) | 4.62 (13.61) | 9.00 (0.28, 17.72) | **0.044** |
| HVLT-R Delayed Recall | 4 ΔBL | 1.75 (10.98) | 0.18 (15.22) | -1.56 (-11.95, 8.83) | 0.758 |
|  | 8 ΔBL | 1.25 (10.47) | 1.44 (14.44) | 0.20 (-10.13, 10.52) | 0.969 |
| D-KEFS Color Naming | 4 ΔBL | 1.80 (5.67) | 5.85 (14.42) | 4.05 (-3.52, 11.62) | 0.282 |
|  | 8 ΔBL | 3.70 (5.44) | 11.10 (14.06) | 7.40 (0.05, 14.75) | **0.049** |
| D-KEFS Word Reading | 4 ΔBL | 3.70 (12.92) | 8.45 (13.28) | 4.75 (-5.84, 15.34) | 0.359 |
|  | 8 ΔBL | 7.90 (12.49) | 9.10 (14.81) | 1.20 (-9.52, 11.92) | 0.818 |
| D-KEFS Inhibition | 4 ΔBL | 7.50 (9.03) | 12.40 (15.08) | 4.90 (-4.17, 13.97) | 0.277 |
|  | 8 ΔBL | 14.30 (11.52) | 14.65 (16.29) | 0.35 (-10.27, 10.97) | 0.946 |
| D-KEFS Inhibition/Switching | 4 ΔBL | 9.10 (7.84) | 9.90 (13.20) | 0.80 (-7.11, 8.71) | 0.837 |
|  | 8 ΔBL | 11.50 (12.28) | 12.65 (13.40) | 1.15 (-9.09, 11.39) | 0.817 |
| D-KEFS Letter Fluency | 4 ΔBL | 2.70 (8.06) | 5.35 (7.98) | 2.65 (-3.88, 9.18) | 0.405 |
|  | 8 ΔBL | 5.00 (6.53) | 6.15 (8.71) | 1.15 (-4.72, 7.02) | 0.689 |
| D-KEFS Category Switching | 4 ΔBL | 1.10 (12.30) | 0.95 (8.64) | -0.15 (-9.49, 9.19) | 0.973 |
|  | 8 ΔBL | 3.60 (7.21) | 5.10 (8.84) | 1.50 (-4.76, 7.76) | 0.624 |
| TMT A | 4 ΔBL | 3.11 (8.94) | 6.45 (10.33) | 3.34 (-4.59, 11.26) | 0.388 |
|  | 8 ΔBL | 5.11 (13.18) | 12.50 (11.69) | 7.39 (-3.58, 18.36) | 0.170 |
| TMT B | 4 ΔBL | 7.11 (6.45) | 6.05 (9.86) | -1.06 (-7.43, 5.31) | 0.734 |
|  | 8 ΔBL | 7.11 (10.22) | 8.00 (9.10) | 0.89 (-7.62, 9.40) | 0.826 |
| Ruff 2&7 Automatic Speed | 4 ΔBL | 2.50 (5.60) | 5.70 (6.39) | 3.20 (-1.54, 7.94) | 0.175 |
|  | 8 ΔBL | 3.90 (7.37) | 9.70 (9.22) | 5.80 (-0.65, 12.25) | 0.076 |
| Ruff 2&7 Controlled Speed | 4 ΔBL | 3.90 (8.33) | 5.75 (9.34) | 1.85 (-5.16, 8.86) | 0.588 |
|  | 8 ΔBL | 3.40 (9.78) | 9.00 (9.94) | 5.60 (-2.39, 13.59) | 0.158 |
| Ruff 2&7 Total Speed | 4 ΔBL | 3.00 (7.07) | 5.95 (7.75) | 2.95 (-2.96, 8.86) | 0.310 |
|  | 8 ΔBL | 3.50 (8.54) | 9.45 (8.94) | 5.95 (-1.08, 12.98) | 0.093 |
| BVMT Total Recall | 4 ΔBL | 5.10 (10.07) | 2.35 (14.06) | -2.75 (-11.98, 6.48) | 0.545 |
|  | 8 ΔBL | -1.67 (19.97) | 1.50 (11.40) | 3.17 (-12.63, 18.96) | 0.666 |
| *Mood and Quality of Life Outcomes* |  |  |  |  |  |
| PHQ-9 | 4 ΔBL | -1.11 (4.59) | -2.47 (5.53) | -1.36 (-5.53, 2.80) | 0.502 |
|  | 8 ΔBL | -2.33 (5.70) | -2.75 (5.16) | -0.42 (-5.18, 4.35) | 0.854 |
| GAD-7 | 4 ΔBL | 0.20 (4.39) | -2.55 (3.10) | -2.75 (-6.09, 0.59) | 0.099 |
| | 8 ΔBL | -0.10 (3.81) | -3.00 (3.32) | -2.90 (-5.90, 0.10) | 0.057 |
| DSQ-PEM | 4 ΔBL | -1.50 (3.12) | -1.74 (8.69) | -0.24 (-4.93, 4.46) | 0.918 |
| | 8 ΔBL | 1.25 (8.14) | -2.32 (6.57) | -3.57 (-10.71, 3.58) | 0.296 |
| SF-36 Emotional Well-Being | 4 ΔBL | 0.40 (15.14) | 7.60 (14.09) | 7.20 (-4.89, 19.29) | 0.226 |
| | 8 ΔBL | -10.00 (20.33) | 9.20 (11.10) | 19.20 (4.15, 34.25) | **0.017** |
| PROMIS Fatigue | 4 ΔBL | -2.68 (6.20) | -2.57 (6.11) | 0.12 (-4.91, 5.14) | 0.962 |
| | 8 ΔBL | 6.10 (7.61) | -2.19 (11.23) | -8.29 (-16.47, -0.11) | **0.047** |

†Two-sided t-test

### Mood and Quality of Life Outcomes

Both groups demonstrated reductions in depressive and anxiety symptoms over time (Table 3). PHQ-9 and GAD-7 scores in the active group showed clinically meaningful improvements, though between-group differences were not statistically significant. Trending improvements in DSQ-PEM scores were observed in the active treatment group. A small effect size was calculated in the GAD-7 (d= -0.428, p= 0.057), and DSQ-PEM (d= -0.338, p= 0.296).

On the SF-36 Emotional Well-Being subscale, participants receiving active MMT showed a significantly greater improvement from baseline to Week 8 compared with sham with a large effect size (d= 0.892, p= 0.017). A significantly greater reduction in fatigue (p= 0.047) was also seen in the active MMT group.

### Usability and User Experience

Participants reported high usability and satisfaction with the device. The majority endorsed that the device was easy to use, comfortable, reliable, and convenient, and most indicated they would recommend it to others (Supplementary Table 1).

## Discussion

This triple-blind, first-in-human, randomized, sham-controlled study demonstrates that at-home MMT is feasible, safe, and well tolerated in individuals with moderate-to-severe cognitive impairment related to LC. Participants showed 100% treatment compliance and high usability ratings, with no device-related adverse events, indicating strong feasibility and an excellent safety profile for at-home implementation.

MMT treatment was also associated with clinically meaningful improvements in cognition and mood at treatment completion and at 8-week follow-up. Several outcomes measuring attention, processing speed, inhibition, and memory significantly improved with active treatment compared to sham treatment. Significant improvements in emotional well-being and clinically meaningful reductions in depressive and anxiety symptoms were also demonstrated in the active group.

While this exploratory study was not powered for efficacy conclusions, consistent improvements across neurocognitive domains, the durability of effect at follow-up, and the clinically meaningful magnitude of change support the potential clinical value of MMT to address an urgent, unmet need in this population.

The magnitude of cognitive improvement observed with MMT compares favorably to other interventions studied in LC populations. Cognitive rehabilitation approaches have demonstrated modest benefits in pilot studies, with small-to-medium effect sizes and improvements largely limited to self-reported cognitive functioning.^16,22,23^ Pharmacologic interventions targeting cognition in LC remain largely untested in rigorous trials, and those repurposed from other conditions (e.g., stimulants, cholinesterase inhibitors) lack evidence specific to post-viral populations. Notably, MMT produced improvements across multiple cognitive measures, including processing speed, attention, and memory, suggesting potential for broader cognitive benefit within the very domains impacted in LC. Furthermore, unlike cognitive rehabilitation protocols that require sustained mental effort and may be limited by post-exertional symptom exacerbation, MMT is passive and low-burden, potentially improving accessibility to patients who cannot tolerate effortful interventions.

### Mechanistic Inferences

The pattern of cognitive improvement observed in this study is consistent with emerging mechanistic models of LC-related cognitive impairment that emphasize persistent neuroinflammation, oxidative stress, and mitochondrial impairment. Improvements in DSS, HVLT-R recall, D-KEFS Color Naming, and SF-36 emotional well-being were strengthened at 8-week follow-up from immediately post-treatment, suggesting persistence of benefit and durability of treatment effects. It is possible that the delay of these effects suggests a mechanism involving gradual resolution of neuroinflammation and restoration of neuronal and glial function, rather than short term expectancy effects alone.

This is the first time that an MMT device has been used in a clinical population, and therefore mechanistic understanding is still emerging. Recent preclinical work found that brief transcranial exposure to MMT suppressed inflammatory signaling in human immune cells, and repeated transcranial MMT attenuated microgliosis, astrogliosis, immune cell infiltration, and tissue degeneration in a rat model of Parkinson’s disease neuroinflammation.^19^ In this work, MMT also reduced neuronal oxidative stress and improved intrinsic antioxidant capacity, with neuroprotective effects persisting for up to 48 hours following a single exposure.^19^ Thus, it is feasible that the clinical gains observed in this study are due to the ability of MMT to suppress neuroinflammation, improve mitochondrial function and promote improved regulation of neurons and glia as these are all mechanisms of LC-related cognitive loss that have been previously described.^11,24–26^

MMT is distinct from conventional neuromodulation approaches in several important ways. The intervention delivers low amplitude, nonthermal radiofrequency magnetic fields to the whole brain and was developed to target neuroinflammatory signaling and cellular metabolic function, rather than directly stimulate or inhibit neuronal firing through focal electrical, magnetic, or ultrasound stimulation. The therapy is designed for self-administration in the home environment using a head worn device. Automated treatment lockout intervals and remote monitoring supported consistent dosing while minimizing user burden and protocol deviation. These characteristics may enable scalable deployment if efficacy is confirmed, particularly for individuals with chronic illness, fatigue related disability, or limited access to in-person therapy.

### Limitations and Future Implications

This study has several limitations. The small sample size limited statistical power, and multiple outcomes were assessed without correction for multiple comparisons. Findings should be interpreted as exploratory.

Repeated neuropsychological testing can introduce practice effects. However, the sham-controlled design helps minimize this influence, and the domain-specific pattern of improvement along with the greater magnitude of change at delayed follow-up compared to immediately post-treatment suggest that the findings are unlikely driven by practice effects alone. Despite these limitations, consistent improvements across neurocognitive domains and mood, the presence of statistically significant and clinically meaningful improvements on certain measures, and the excellent feasibility and safety profile provide a strong rationale for future investigation. These findings may inform the design of larger clinical trials to confirm efficacy and clarify mechanisms.

## Conclusions

MMT represents a promising, nonpharmacologic intervention for cognitive impairment associated with LC. This study supports the feasibility and safety of MMT and provides preliminary evidence of sustained cognitive improvements and benefits in mood-related symptoms in people with LC.

## Data Availability

All data produced in the present study are available upon reasonable request to the authors.

## References

1. Mandel H, Yoo YJ, Allen AJ, et al. Long COVID Incidence Proportion in Adults and Children Between 2020 and 2024: An Electronic Health Record-Based Study From the RECOVER Initiative. Clinical Infectious Diseases. 2025;80(6):1247–1261. doi:10.1093/cid/ciaf046

2. Goldowitz I, Worku T, Brown L, Fineberg H V. A Long COVID Definition: A Chronic, Systemic Disease State with Profound Consequences. National Academies Press (US); 2024. doi:10.17226/27768

3. Ford ND, Agedew A, Dalton AF, Singleton J, Perrine CG, Saydah S. Notes from the Field: Long COVID Prevalence Among Adults — United States, 2022. MMWR Morb Mortal Wkly Rep. 2024;73(6):135-135–136. 10.15585/mmwr.mm7306a4

4. Proal AD, VanElzakker MB, Aleman S, et al. SARS-CoV-2 reservoir in post-acute sequelae of COVID-19 (PASC). Nat Immunol. 2023;24(10):1616–1627. doi:10.1038/s41590-023-01601-2

5. Iwasaki A, Putrino D. Why we need a deeper understanding of the pathophysiology of long COVID. Lancet Infect Dis. 2023;23(4):393–395. doi:10.1016/S1473-3099(23)00053-1

6. Davis HE, McCorkell L, Vogel JM, Topol EJ. Long COVID: major findings, mechanisms and recommendations. Nat Rev Microbiol. 2023;21(3):133–146. doi:10.1038/s41579-022-00846-2

7. Cutler DM. The Costs of Long COVID. JAMA Health Forum. 2022;3(5):e221809–e221809. doi:10.1001/jamahealthforum.2022.1809

8. Ceban F, Ling S, Lui LMW, et al. Fatigue and cognitive impairment in Post-COVID-19 Syndrome: A systematic review and meta-analysis. Brain Behav Immun. 2022;101:93–135. 10.1016/j.bbi.2021.12.020

9. Becker JH, Lin JJ, Doernberg M, et al. Assessment of Cognitive Function in Patients After COVID-19 Infection. JAMA Netw Open. 2021;4(10):e2130645–e2130645. doi:10.1001/jamanetworkopen.2021.30645

10. VanElzakker MB, Bues HF, Brusaferri L, et al. Neuroinflammation in post-acute sequelae of COVID-19 (PASC) as assessed by [11C]PBR28 PET correlates with vascular disease measures. Brain Behav Immun. 2024;119:713–723. 10.1016/j.bbi.2024.04.015

11. Fernández-Castañeda A, Lu P, Geraghty AC, et al. Mild respiratory COVID can cause multi-lineage neural cell and myelin dysregulation. Cell. 2022;185(14):2452–2468.e16. doi:10.1016/j.cell.2022.06.008

12. Stefano GB, Ptacek R, Ptackova H, Martin A, Kream RM. Selective neuronal mitochondrial targeting in SARS-CoV-2 infection affects cognitive processes to induce “brain fog” and results in behavioral changes that favor viral survival. Med Sci Monit. 2021;27:e930886. doi:10.12659/MSM.930886

13. Braga J, Lepra M, Kish SJ, et al. Neuroinflammation After COVID-19 With Persistent Depressive and Cognitive Symptoms. JAMA Psychiatry. 2023;80(8):787–795. doi:10.1001/jamapsychiatry.2023.1321

14. Visser D, Golla SS V, Verfaillie SCJ, et al. Long COVID is associated with extensive *in-vivo* neuroinflammation on [^18^F]DPA-714 PET. medRxiv. Published online January 1, 2022:2022.06.02.22275916. doi:10.1101/2022.06.02.22275916

15. Becker JH, Lin JJ, Twumasi A, et al. Greater executive dysfunction in patients post-COVID-19 compared to those not infected. Brain Behav Immun. 2023;114:111–117. 10.1016/j.bbi.2023.08.014

16. Becker JH, Li J, Lin JJ, et al. Neurocognitive trajectories in long COVID: Evidence from longitudinal analyses. Brain Behav Immun Health. 2025;48:101093. 10.1016/j.bbih.2025.101093

17. Rohde CH, Taylor EM, Alonso A, Ascherman JA, Hardy KL, Pilla AA. Pulsed Electromagnetic Fields Reduce Postoperative Interleukin-1β, Pain, and Inflammation: A Double-Blind, Placebo-Controlled Study in TRAM Flap Breast Reconstruction Patients. Plast Reconstr Surg. 2015;135(5). https://journals.lww.com/plasreconsurg/fulltext/2015/05000/pulsed_electromagnetic_field s_reduce_postoperative.4.aspx

18. Rasouli J, Lekhraj R, White NM, et al. Attenuation of interleukin-1beta by pulsed electromagnetic fields after traumatic brain injury. Neurosci Lett. 2012;519(1):4–8. 10.1016/j.neulet.2012.03.089

19. Nguyen N, Brady NR, Timblin GA, Tharp KM, Gurfein BT. Transcranial microtesla magnetic fields suppress neuroinflammation and neuronal oxidative stress burden. iScience. 2026;29(1):114425. 10.1016/j.isci.2025.114425

20. Muir RT, Hill MD, Black SE, Smith EE. Minimal clinically important difference in Alzheimer’s disease: Rapid review. Alzheimer’s & Dementia. 2024;20(5):3352–3363. 10.1002/alz.13770

21. Kounali D, Button KS, Lewis G, et al. How much change is enough? Evidence from a longitudinal study on depression in UK primary care. Psychol Med. 2022;52(10):1875–1882. doi:DOI: 10.1017/S0033291720003700

22. Herrera E, Pérez-Sánchez M del C, San Miguel-Abella R, et al. Cognitive impairment in young adults with post COVID-19 syndrome. Sci Rep. 2023;13(1):6378. doi:10.1038/s41598-023-32939-0

23. Möller M, Borg K, Janson C, Lerm M, Normark J, Niward K. Cognitive dysfunction in post-COVID-19 condition: Mechanisms, management, and rehabilitation. J Intern Med. 2023;294(5):563–581. doi:10.1111/joim.13720

24. Matits L, Schellenberg J, Mack M, et al. Circulating mitochondrial and cellular damage markers in long COVID: Links to cognitive function, psychological distress, and inflammation. Mol Psychiatry. Published online 2026. doi:10.1038/s41380-026-03471-0

25. Szögi T, Borsos BN, Masic D, et al. Novel biomarkers of mitochondrial dysfunction in Long COVID patients. Geroscience. 2025;47(2):2245–2261. doi:10.1007/s11357-024-01398-4

26. Xu W tao, An X bin, Chen M jie, et al. A Gene Cluster of Mitochondrial Complexes Contributes to the Cognitive Decline of COVID-19 Infection. Mol Neurobiol. 2025;62(6):6869–6883. doi:10.1007/s12035-024-04471-3

