## Supplementary Table 1 for "Microtesla Magnetic Therapy for cognitive impairment in post-acute sequelae of SARS CoV-2: A randomized controlled feasibility study"

Supplementary Table 1. Counts of Participant Responses to Device Usability Ratings (N = 30)

| **Question** | **1 - Disagree** | **2** | **3** | **4** | **5 - Agree** |
| --- | --- | --- | --- | --- | --- |
| I always knew how much time remained in a therapy session. | 3 | 0 | 0 | 4 | 23 |
| I can rely on the study device to deliver therapy consistently. | 0 | 1 | 4 | 4 | 21 |
| I found the audio indicating therapy helpful. | 0 | 0 | 2 | 7 | 21 |
| I would recommend the study device to others. | 0 | 0 | 6 | 4 | 20 |
| The controls of study device were easy to understand. | 0 | 0 | 2 | 2 | 26 |
| The device instructions were easy to understand. | 0 | 0 | 1 | 5 | 24 |
| The study device was comfortable to wear during therapy. | 0 | 0 | 3 | 4 | 23 |
| The study device was easy to use. | 0 | 0 | 0 | 5 | 25 |
| Therapy with the study device was convenient. | 0 | 0 | 0 | 5 | 25 |
