## Supplementary Table 2 for "Microtesla Magnetic Therapy for cognitive impairment in post-acute sequelae of SARS CoV-2: A randomized controlled feasibility study"

Supplementary Table 2. Effect Size for Outcomes

| **Measure** | **Week** | **Cohen's d** | **Effect Size** |
| --- | --- | --- | --- |
| HVLT-R Recall | 0 | -0.540 | medium |
| HVLT-R Recall | 4 | -0.067 |  |
| HVLT-R Recall | 8 | 0.229 | small |
| Digit Span Forward | 0 | 0.124 |  |
| Digit Span Forward | 4 | 0.563 | medium |
| Digit Span Forward | 8 | 0.225 | small |
| Digit Span Backward | 0 | 0.042 |  |
| Digit Span Backward | 4 | -0.046 |  |
| Digit Span Backward | 8 | 0.316 | small |
| Digit Span Sequencing | 0 | -0.399 | small |
| Digit Span Sequencing | 4 | 0.087 |  |
| Digit Span Sequencing | 8 | 0.536 | medium |
| D-KEFS Color Naming | 0 | -0.151 |  |
| D-KEFS Color Naming | 4 | 0.210 | small |
| D-KEFS Color Naming | 8 | 0.698 | medium |
| D-KEFS Inhibition | 0 | 0.099 |  |
| D-KEFS Inhibition | 4 | 0.569 | medium |
| D-KEFS Inhibition | 8 | 0.173 |  |
| D-KEFS Letter Fluency | 0 | 0.220 | small |
| D-KEFS Letter Fluency | 4 | 0.483 | small |
| D-KEFS Letter Fluency | 8 | 0.327 | small |
| D-KEFS Word Reading | 0 | 0.295 | small |
| D-KEFS Word Reading | 4 | 1.078 | large |
| D-KEFS Word Reading | 8 | 0.763 | medium |
| TMT A | 0 | 0.061 |  |
| TMT A | 4 | 0.609 | medium |
| TMT A | 8 | 0.664 | medium |
| Ruff 2 & 7 Auto Speed | 0 | 0.193 |  |
| Ruff 2 & 7 Auto Speed | 4 | 0.413 | small |
| Ruff 2 & 7 Auto Speed | 8 | 0.574 | medium |
| Ruff 2 & 7 Controlled Speed | 0 | 0.314 | small |
| Ruff 2 & 7 Controlled Speed | 4 | 0.390 | small |
| Ruff 2 & 7 Controlled Speed | 8 | 0.615 | medium |
| Ruff 2 & 7 Total Speed | 0 | 0.267 | small |
| Ruff 2 & 7 Total Speed | 4 | 0.442 | small |
| Ruff 2 & 7 Total Speed | 8 | 0.619 | medium |
| Symbol Digit Modalities Test | 0 | 0.069 |  |
| Symbol Digit Modalities Test | 4 | 0.676 | medium |
| Symbol Digit Modalities Test | 8 | -0.081 |  |
| PHQ-9 | 0 | -0.022 |  |
| PHQ-9 | 4 | -0.360 | small |
| PHQ-9 | 8 | -0.161 |  |
| GAD-7 | 0 | 0.132 |  |
| GAD-7 | 4 | -0.495 | small |
| GAD-7 | 8 | -0.428 | small |
| DSQ-PEM | 0 | -0.081 |  |
| DSQ-PEM | 4 | -0.077 |  |
| DSQ-PEM | 8 | -0.338 | small |
| SF-36 Emotional Wellbeing | 0 | -0.127 |  |
| SF-36 Emotional Wellbeing | 4 | 0.281 | small |
| SF-36 Emotional Wellbeing | 8 | 0.892 | large |
