## Supplementary Table 3 for "Microtesla Magnetic Therapy for cognitive impairment in post-acute sequelae of SARS CoV-2: A randomized controlled feasibility study"

Supplementary Table 3. Between-group comparisons of unimproved outcomes and SF-36 domain scores

| **Outcome** | **Week** | **Sham Mean (SD)** | **Active Mean (SD)** | **Active-Sham Difference (95% CI)** | **P-value†** |
| --- | --- | --- | --- | --- | --- |
| D-KEFS Category Fluency | 4 ∆BL | 1.90 (11.65) | 3.60 (8.57) | 1.70 (-7.20, 10.60) | 0.688 |
|  | 8 ∆BL | 2.90 (6.85) | -0.85 (12.52) | -3.75 (-11.01, 3.51) | 0.299 |
| RCFT | 4 ∆BL | 3.50 (12.09) | 1.58 (10.38) | -1.92 (-11.46, 7.62) | 0.675 |
|  | 8 ∆BL | 0.90 (10.09) | 0.79 (12.93) | -0.11 (-9.13, 8.91) | 0.980 |
| SDMT | 4 ∆BL | -1.80 (12.79) | 4.50 (9.69) | 6.30 (-3.52, 16.12) | 0.191 |
|  | 8 ∆BL | 7.10 (6.33) | 5.35 (9.57) | -1.75 (-7.78, 4.28) | 0.556 |
| MINT | 4 ∆BL | -0.40 (5.02) | 1.30 (8.77) | 1.70 (-3.47, 6.87) | 0.506 |
|  | 8 ∆BL | 5.90 (5.70) | 2.50 (6.66) | -3.40 (-8.27, 1.47) | 0.161 |
| SF-36 Energy / Fatigue | 4 ∆BL | 5.50 (21.01) | 4.25 (13.60) | -1.25 (-17.05, 14.55) | 0.867 |
|  | 8 ∆BL | 1.00 (9.94) | 2.00 (16.89) | 1.00 (-9.08, 11.08) | 0.840 |
| SF-36 General Health | 4 ∆BL | -4.50 (13.63) | 0.25 (14.73) | 4.75 (-6.59, 16.09) | 0.392 |
|  | 8 ∆BL | 0.00 (8.16) | -1.00 (12.94) | -1.00 (-8.97, 6.97) | 0.799 |
| SF-36 Pain | 4 ∆BL | 5.75 (15.09) | 0.63 (13.13) | -5.13 (-17.00, 6.75) | 0.374 |
|  | 8 ∆BL | 7.75 (20.80) | -3.38 (21.29) | -11.13 (-28.15, 5.90) | 0.187 |
| SF-36 Physical Functioning | 4 ∆BL | 1.00 (18.68) | -0.25 (11.64) | -1.25 (-15.24, 12.74) | 0.850 |
|  | 8 ∆BL | -13.00 (43.60) | 0.75 (12.70) | 13.75 (-17.72, 45.22) | 0.352 |
| SF-36 Role Limitations - Emotional | 4 ∆BL | 13.33 (61.26) | 5.00 (47.48) | -8.33 (-55.53, 38.87) | 0.711 |
|  | 8 ∆BL | 30.00 (59.73) | 1.67 (48.94) | -28.33 (-74.80, 18.13) | 0.214 |
| SF-36 Role Limitations - Physical | 4 ∆BL | 17.50 (31.29) | 12.50 (37.61) | -5.00 (-31.98, 21.98) | 0.704 |
|  | 8 ∆BL | 15.00 (29.34) | -7.50 (28.21) | -22.50 (-46.13, 1.13) | 0.061 |
| SF-36 Social Functioning | 4 ∆BL | -3.75 (29.49) | 0.00 (14.05) | 3.75 (-17.89, 25.39) | 0.710 |
|  | 8 ∆BL | 5.00 (22.97) | -1.25 (18.09) | -6.25 (-24.00, 11.50) | 0.464 |
| SF-36 Total Score | 4 ∆BL | 35.23 (104.27) | 29.98 (92.63) | -5.26 (-87.68, 77.17) | 0.894 |
|  | 8 ∆BL | 35.75 (146.79) | 0.49 (102.41) | -35.26 (-146.64, 76.12) | 0.507 |
